# Single-cell RNA sequencing unraveled the expression heterogeneity of hematopoietic stem and progenitor cells and lymphoid cell development dysregulation in childhood asthma

**DOI:** 10.1101/2024.03.17.24304334

**Authors:** Danying Zhu, Guang Li, Lang Yuan, Zeyu Zeng, Na Dong, Chao Wang, Ming Chen, Lijian Xie, Guohui Ding, Libing Shen, Xiaoyan Dong

## Abstract

Asthma is a long-term inflammatory disease affecting airways and lungs with usual onset in childhood. Its cause is not fully understood up to now. Here, using single-cell RNA sequencing, we profile peripheral blood mononuclear cells (PBMCs) from three pediatric patients with onset asthma and four age-matched healthy controls to investigate the cellular etiology of childhood asthma. The overall expression features among three asthma patients’ PBMCs demonstrate that innate immunity is commonly upregulated while adaptive immunity is commonly downregulated in childhood asthma, but each patient has different molecular phenotypes. The analyses of the expression profiles of hematopoietic stem and progenitor cells (HSPCs) further show that the HSPCs of asthma patients have heterogeneous expression backgrounds with more specific differentially expressed genes (DEGs) in each patient than common DEGs and a common feature of low S100 protein binding gene expression. S100A8, S100A9, S100A12, and RETN are universally upregulated in various cell types of asthma patients. The cell developmental trajectories in three asthma cases exhibit an abnormal immune cell development pattern compared to that in health control. The dysregulated lymphoid lineage development is observed in all 3 patients, but there is no identical abnormal pattern for each patient. The pseudo-time analyses of gene expression show that the expression dynamics of two proto-oncogenes, JUN and SPI1, and six inflammatory response related genes (S100A8, S100A9, S100A12, IL7R, IL32, CCL5) are relevant to abnormal immune cell development in asthma patients. The cell-cell communication analyses reveal the contribution of incoming annexin signal towards dendritic cells and the outgoing resistin signal from dendritic cells to asthma heterogeneity. Interestingly, the plasma blast cells of asthma patient 3 with severe symptoms exhibit dual cell identities of both plasma blast cells and T cells. Our scRNA-Seq analyses for three asthma patients reveal a complex cellular etiology for childhood asthma and provide a new research direction for the comprehensive and systematic understanding of key molecular mechanisms of childhood asthma.

## Introduction

Asthma, a persistent respiratory condition, is marked by inflammation of the airways, heightened bronchial responsiveness, and variable airway obstruction. This complex disease involves a multitude of cellular participants, including eosinophils, T and B lymphocytes, macrophages, neutrophils, and epithelial cells, which interact with environmental factors to induce chronic inflammation, reversible airflow limitations, and airway remodeling in patients^1^. The involvement of various cell types and their mediators is a key aspect of asthma’s pathogenesis^2^. This diversity in asthma phenotypes and the intricacy of its pathogenesis are further complicated by the fact that respiratory infections and allergen exposures are known to trigger asthma exacerbations^1^. While asthma can affect individuals across all age groups, the manifestation, underlying pathophysiology, and response to treatment can vary significantly between children and adults. Recognizing the molecular and cellular distinctions between childhood and adult asthma is crucial for crafting age-tailored diagnostic and therapeutic approaches.

childhood asthma, in particular, presents with distinct clinical and immunological characteristics compared to its adult counterpart. Asthma in children often manifests as allergic phenotypes with a higher incidence of eosinophilic inflammation, whereas adults may display a broader range of subtypes, including non-eosinophilic variants. The immature and plastic nature of the immune system in children can lead to unique cellular responses and immune dysregulation. Therefore, there are some differences in the treatment by unique age groups^3^. Single-cell sequencing offers a powerful tool to explore age-specific alterations in immune cell populations, their activation states, and functional attributes, providing a deeper understanding of the mechanisms driving childhood asthma.

Previous research has predominantly focused on the single-cell sequencing of adult asthma^4, 5^, whereas this study applies single-cell technology to childhood asthma, aiming to uncover the role of immune abnormalities in the pathogenesis of asthma in children. Through single-cell sequencing, specific cell subpopulations can be identified, and their roles in the inflammatory responses of children with asthma can be understood, as well as their responses to current treatment methods. These findings may help explain why childhood asthma exhibits differences in clinical presentation and treatment response compared to adult asthma, and they provide new perspectives for future research and clinical practice.

## Method

### Patients

During the initial hospital stay, before any treatment, we collected three fresh peripheral blood samples from patients experiencing acute asthma. The diagnosis of asthma was confirmed using the criteria outlined in the 2023 Global Initiative for Asthma (GINA) report, which can be accessed at https://ginasthma.org/2023-gina-main-report/. Atopic status was determined by a total IgE level of 200 IU/ml or higher. For single-cell RNA sequencing (scRNA-seq), we enrolled four healthy individuals who were participating in routine physical examinations and had no recent history of fever, infection, or immunization. Data from three of these individuals have previously been published^6^. All participants were recruited from Shanghai Children’s Hospital between December 2019 and February 2023. Exclusion criteria for the study included: Participants who were undergoing immunotherapy, such as anti-IgE treatment, or had received such therapy within the last three months. Individuals with significant abnormalities identified in their complete blood count. Subjects who were currently enrolled in an asthma-related pharmaceutical or interventional study, or had been part of such a study within the last four weeks. Patients with concurrent medical conditions necessitate systemic corticosteroids or other immunomodulatory treatments. The study was approved by the Ethics Committee of Shanghai Children’s Hospital (Protocol Numbers: 2019R081, 2022R029-F-01). Informed consent was obtained from all participants and their guardians.

### Single-cell preparation and sequencing

A total of 2 milliliters of venous blood was collected from each participant using tubes containing EDTA as an anticoagulant. These samples were processed within four hours to preserve cell integrity. Peripheral blood mononuclear cells (PBMCs) were separated from the collected blood using Ficoll-Paque medium and density gradient centrifugation. The viability of the isolated PBMCs was assessed using trypan blue staining, and only samples with more than 90% viable cells were selected for further analysis. A calculated volume of the cell suspension, estimated to contain around 12,000 cells per sample, was prepared to ensure an adequate cell count for subsequent capture and sequencing processes. The Chromium Next GEM Single Cell V(D)J Reagent Kits v1.1 from 10x Genomics were utilized for capturing single cells and constructing the corresponding libraries. The cell suspension, along with barcoded gel beads and partitioning oil, were loaded onto the 10 × Genomics Chromium Chip to create single-cell Gel Beads-in-Emulsion (GEMs). Inside each GEM, cells were lysed, and their transcripts were barcoded through reverse transcription. After reverse transcription, the cDNA, now carrying cell barcodes, underwent PCR amplification to ensure a sufficient quantity for sequencing. The construction of single-cell RNA sequencing (scRNA-seq) libraries was accomplished using the 5’ Library Kits. The prepared libraries were sequenced on an Illumina NovaSeq platform, producing paired-end reads with a length of 2 × 150 base pairs. This sequencing step generates the raw data necessary for subsequent data analysis.

### scRNA-seq data analysis

The demultiplexed reads were then aligned to the GRCh38 reference genome using the Cell Ranger software suite. This gene-barcode matrix was subsequently analyzed using Seurat version 3.0.2 for various analytical steps, including quality control, data normalization, dimensionality reduction, batch effect correction, clustering, and data visualization. The specific procedures carried out with Seurat included: Applying quality control criteria for the majority of samples, which involved ensuring a total UMI count ranging from 200 to 6000 and limiting the percentages of mitochondrial, hemoglobin, and ribosome genes to below 10%, 0.1%, and 3%, respectively. Integrating samples collected during the initial phase of the study (C1-C4) to eliminate batch effects using anchor. Scaling the integrated matrix and employing the principal components derived from principal component analysis (PCA) for uniform manifold approximation and projection (UMAP). Utilizing UMAP to visualize cells in a two-dimensional space, highlighted their gene expression pattern similarities. Conducting shared nearest neighbor graph-based clustering on the PCA-reduced data to identify the primary cell types within the PBMCs. Examining the expression of canonical marker genes to refine cell cluster annotations. Determining cell identities through multimodal reference mapping with the SeuratDisk package.

### Differential expression, functional enrichment analysis, and pseudo-time analysis

The marker genes for each cluster were calculated with the FindAllMarkers function within the Seurat package. The clusterProfiler package (version 3.16.0) was used for function over-representation analysis of the differentially expressed genes (DEGs) with a false discovery rate (FDR) threshold of <0.05. Gene Ontology (GO), KEGG pathways, and hallmark gene sets from the MSigDB database (version 7.1) were used as gene function databases to assess the enrichment of specific biological functions or pathways among the DEGs. Pseudo-time analysis of cell differentiation trajectories for each sample dataset was performed with R package Monocle 2^7^. The expression feature and inferred cell type for each sample dataset from the Seurat result were used to annotate the cell dataset for the Monocle analysis pipeline. We used the Monocle built-in approach named “dpFeature” to detect the variable genes that define a cell’s differentiation. Its advantages are needing no prior biological knowledge and discovering important ordering genes from the data itself. Dimension reduction was performed with 2 max components and the “DDRTree” method. Cell-cell communication analysis was performed with the R package CellChat 1.6.1^8^.

### Human serum resistin Enzyme-Linked Immunosorbent Assay (ELISA) analysis

EDTA-anticoagulated whole blood was transferred to the laboratory and processed immediately after collection. Centrifuge samples for 15 minutes at 1000 x g at 4 □ within 30 minutes of collection and stored at −80 □. We followed a human resistin ELISA kit (Signalway Antibody, Maryland, USA, Catalog No: EK2351) protocol to analyze peripheral blood serums.

## Results

### Characteristics of the study subjects for asthma patients and health controls

In our single-cell RNA analysis study, we included a cohort of 7 childhood subjects, comprising 3 children experiencing asthma exacerbations and 4 healthy individuals without asthma. T The mean age of the participants ranges from 1 to 9 years old. An additional group of 14 asthma patients and health controls ranging from 1 to 13 years old, were included in the study. Allergic testing in asthmatic children revealed sensitization to house dust mites. Allergic testing in asthmatic children revealed sensitization to house dust mites.

### Single-cell RNA profiling of PBMCs in health and asthma patients

We collected the peripheral blood samples derived from 3 patients with onset asthma (A1-A3). The patients were diagnosed according to the GINA 2023 (https://ginasthma.org/2023-gina-main-report/). For each patient, the blood sample was taken on the first days before therapy. We also collected fresh peripheral blood samples from four age-matched healthy donors as controls (C1–C4).

We used the 10× Genomics platform for scRNA-seq of PBMCs isolated from the samples. PBMCs were loaded onto the platform and about 5000 - 14000 cells per sample could be recovered from the sequencing data. The total number of detected cells passing quality control was 34,354 for 3 asthma patients and 22,398 cells from healthy controls. Based on the scRNA-seq profiles, we clustered the cells across samples with Seurat 4.0 and visualized them in two-dimensional space (Fig. 1a). The PBMCs in our study could be clustered into 12 major cell types including B cells, CD4 T cells, CD8 T cells, CD14 monocytes (CD14 mono), CD16 monocytes (CD16 mono), dendritic cells (DC), mucosa-associated invariant T cells (MAIT), gamma-delta cells (gdT), hematopoietic stem and progenitor cells (HSPC), natural killer cells (NK), plasma blast cells (Plasmablast), and platelets. Each cell type was verified with PBMC multimodal reference object and canonical gene markers (Supplementary Figure 1a and 1b). The percentage of major cell types for each sample is shown in Figure 1b. The more detailed classification of cell types excluding CD4 T and CD8 T cells shows that CD14 monocytes are significantly increased and CD16 monocytes are significantly decreased in asthma patients (Figure 1c, *P*-value < 0.05). The detailed classification of T cell types in each sample is shown in Supplementary Figure 1c.

**Figure 1.**
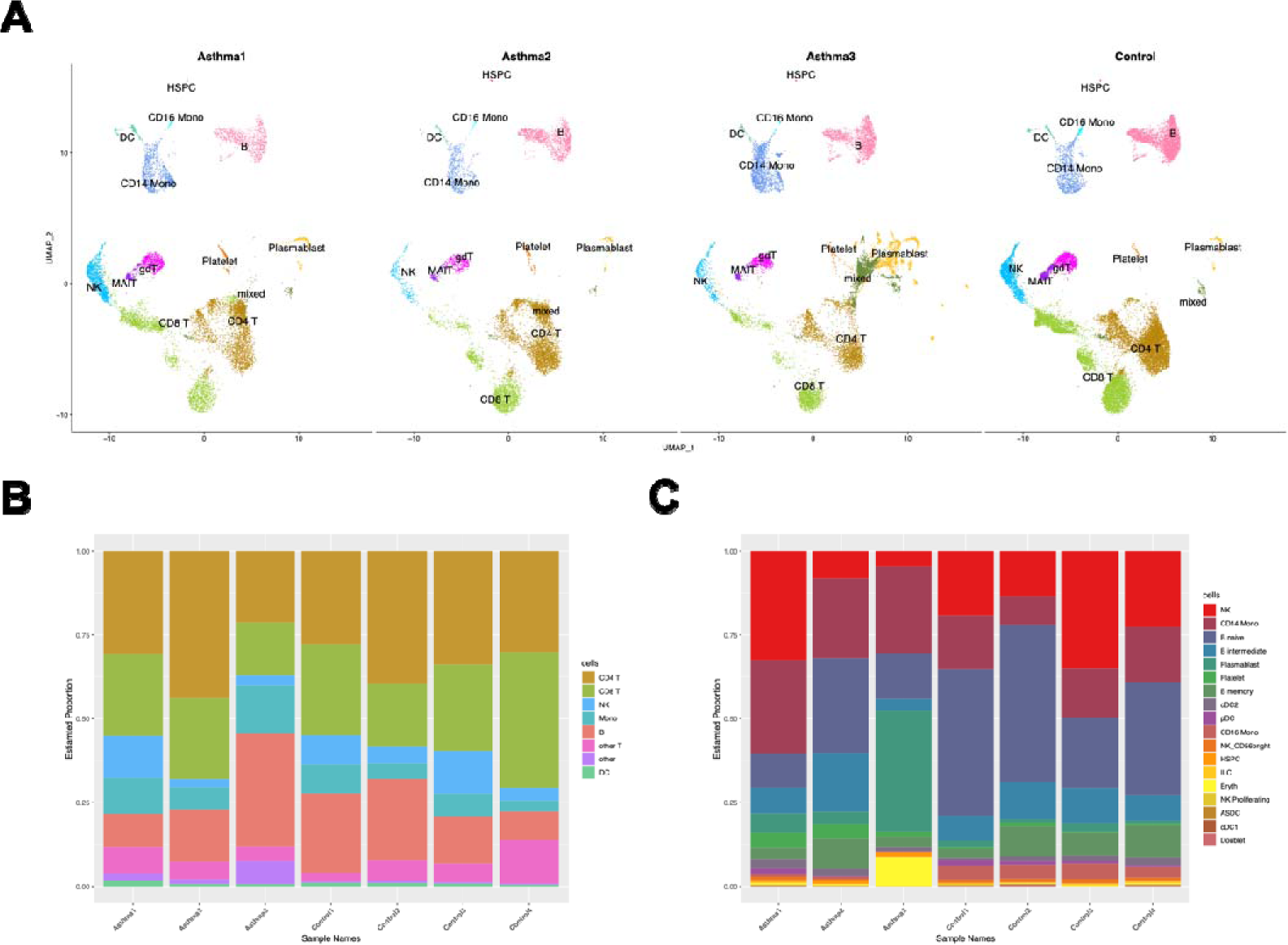
Single-cell profiling of PBMCs in six samples. A. The integration single-cell profiling analysis of six samples including 4 healthy controls, asthma patient 1, asthma patient 2, and asthma patient 3. B. The percentage bar chart shows the continent of different cell types in each sample. C. The percentage bar chart shows the different cell types except T cells in each sample.

### Expression features of all cells and HSPCs in asthma patients

We first compared the gene expression level of all cells in each asthma patient to controls. The overall differentially expressed genes (DEGs) were identified for each asthma patient. There are 174 upregulated genes in A1 (Asthma1), 29 upregulated genes in A2 (Asthma2), and 246 upregulated genes in A3 (Asthma3) (Figure 2a). There are 21 common upregulated genes among three asthma patients. GO analysis shows that these commonly upregulated genes are involved in the extracellular region, secretion, and antimicrobial function (Figure 2b). GO result also shows that innate immunity plays a major role in childhood asthma which manifests as a defense response to bacterium and inflammatory response. There are 57 specific upregulated genes in A1, 3 specific upregulated genes in A2, and 128 specifically upregulated genes in A3. GO analyses of the specifically upregulated genes in A1 and A3 show that these two patients have different molecular phenotypes (Supplemental Tables 1 and 2). A1 exhibits the specific expression of signal sequences (usually 20-30 amino acids long) and positive regulation of T cell migration while A3 has an excessive expression of immunoglobulin. 3 specific upregulated genes in A2 are IGHV3-73, CDK6 and NELL2 which are not sufficient for GO analysis. We also find 33 downregulated genes in A1 (Asthma1), 80 downregulated genes in A2 (Asthma2), and 66 downregulated genes in A3 (Asthma3) (Figure 2c). There are 9 common upregulated genes among three asthma patients. GO analysis shows that these common downregulated genes are involved in adaptive immunity and T cell receptor (Figure 2d). There are 13 specific downregulated genes in A1, 37 specific downregulated genes in A2, and 20 specific downregulated genes in A3. A1 exhibits the specific repression of the external side of the plasma membrane and B cell receptor signaling pathway expression (Supplemental Table 3). A2 exhibits the specific repression of the immunoglobulin complex (Supplemental Table 4). A3 exhibits the specific repression of the T cell receptor complex (Supplemental Table 5).

**Figure 2.**
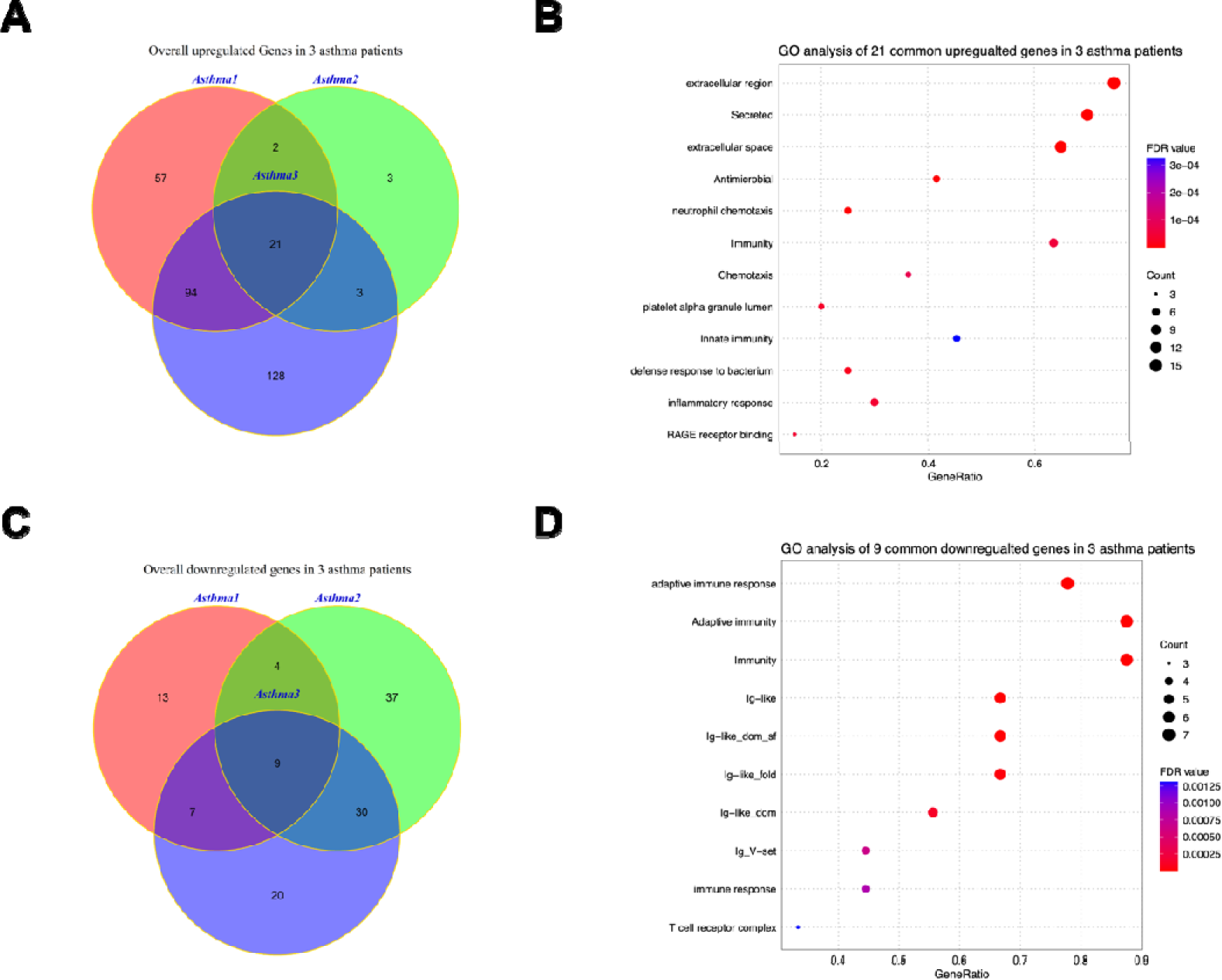
The analysis of upregulated and downregulated genes in asthma patients. A. Venn gram of the upregulated genes in 3 asthma patients. B. GO term enrichment analysis of the specific upregulated genes in 3 asthma patients. C. Venn gram of the downregulated genes in 3 asthma patients. D. GO term enrichment analysis of the specific downregulated genes in 3 asthma patients.

We further examined the expression features of HSPCs in asthma patients, because the other types of cells are all derived from them. There are 176 upregulated HSPC genes in A1 (Asthma1), 211 upregulated HSPC genes in A2 (Asthma2), and 199 HSPC upregulated genes in A3 (Asthma3) (Figure 3a). There are 81 common upregulated HSPC genes among three asthma patients. GO analysis shows that these commonly upregulated HSPC genes are mainly involved in the cellular components of extracellular exosome and ER to Golgi transport vesicle membrane (Figure 3b). There are 29 specifically upregulated HSPC genes in A1, 72 specifically upregulated HSPC genes in A2, and 55 specifically upregulated HSPC genes in A3. No significant GO result is found for 29 specifically upregulated HSPC genes in A1. HSPCs in A2 exhibit a specific expression of extracellular exosome related genes and HSPCs in A3 exhibit a specific expression of endoplasmic reticulum related genes (Supplemental Tables 6 and 7). There are 179 downregulated HSPC genes found in A1 (Asthma1), 203 downregulated HSPC genes found in A2 (Asthma2), and 119 HSPC downregulated genes found in A3 (Asthma3) (Figure 3c). GO analysis shows that these common downregulated HSPC genes are mainly involved in S100 protein binding (Figure 3d). There are 85 specifically downregulated HSPC genes in A1, 113 specifically downregulated HSPC genes in A2, and 53 specifically downregulated HSPC genes in A3. HSPCs in A1 exhibit a specific repression of adaptive immunity and T cell receptor complex expression (Supplemental Table 8). HSPCs in A2 exhibit a specific repression of immune response related genes’ expression (Supplemental Table 9). HSPCs in A3 exhibit a specific repression of extracellular exosome and immunity relate genes’ expression (Supplemental Table 10). Interestingly, we find that T cell receptor genes are repressed in all three asthma patients, although they are not statically significant in the HSPCs of A2 and A3 (Supplemental Tables 9 and 10).

**Figure 3.**
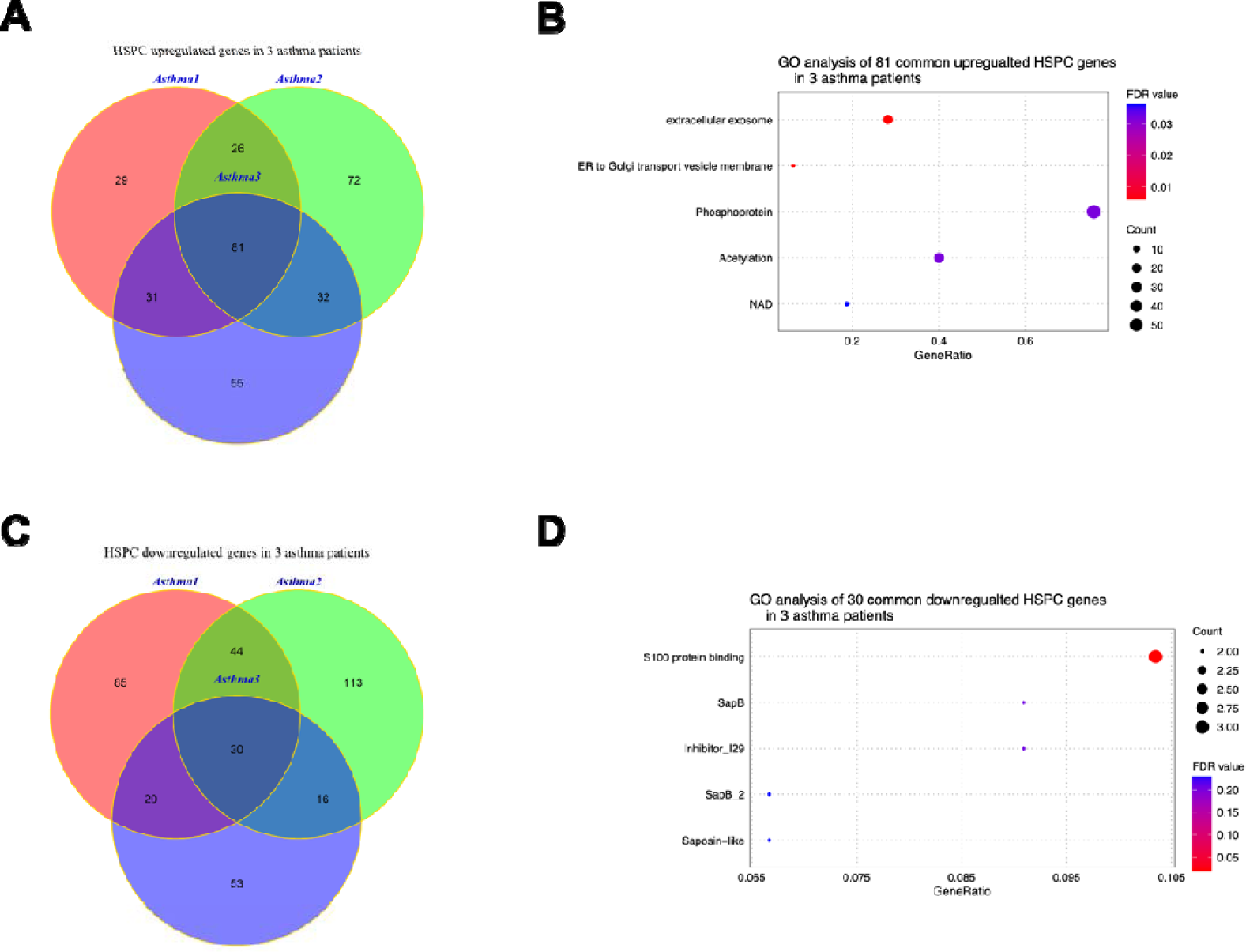
The analysis of upregulated and downregulated genes in asthma patients’ HSPC. A. Venn gram of the upregulated genes in 3 asthma patients’ HSPC. B. GO term enrichment analysis of the specific upregulated genes in 3 asthma patients’ HSPC. C. Venn gram of the downregulated genes in 3 asthma patients’ HSPC. D. GO term enrichment analysis of the specific downregulated genes in 3 asthma patients’ HSPC.

In expression feature analyses, we find that S100A8, S100A9, S100A12, and RETN are universally upregulated in various cell types of asthma patients, especially S100A8 and S100A9 (Figure 4). S100A8, S100A9, and S100A12 are known for participating in inflammatory response while RETN has antibacterial activity^9–11^.

**Figure 4.**
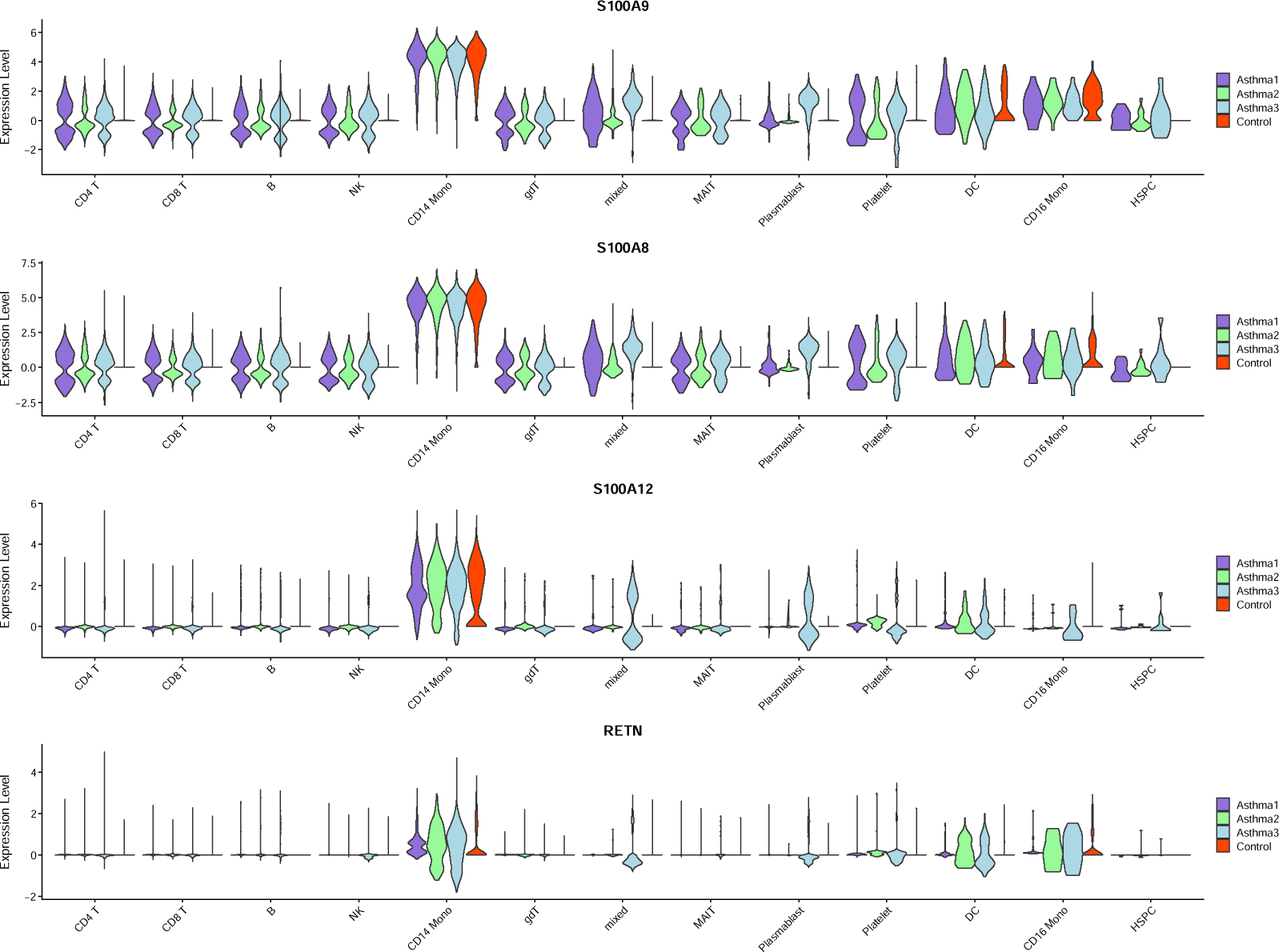
The violin plot of S100A9, S100A8, S100A12 and RETN gene expression levels across different cell types for four sample groups.

### Single-cell trajectory reconstructions of PBMCs in health controls and 3 asthma patients

The expression features of asthma patients’ HSPCs demonstrate that the expression of immunity and T cell receptor genes are abnormally repressed in them. Since HSPCs give rise to the other cells, it is worth investigating the cell developmental states in asthma patients. Single-cell trajectory analysis could help us reconstruct the cell developmental paths of PBMCs based on featured variable genes. The analyses show that the PBMCs in healthy controls can be divided into 5 developmental states while the PBMCs in asthma patients have 7, 3, and 5 developmental states, differently (Figure 5a, 5c, 5e, and 5g). All cell types are plotted on each trajectory in each sample (Figure 5b, 5d, 5f, and 5h).

**Figure 5.**
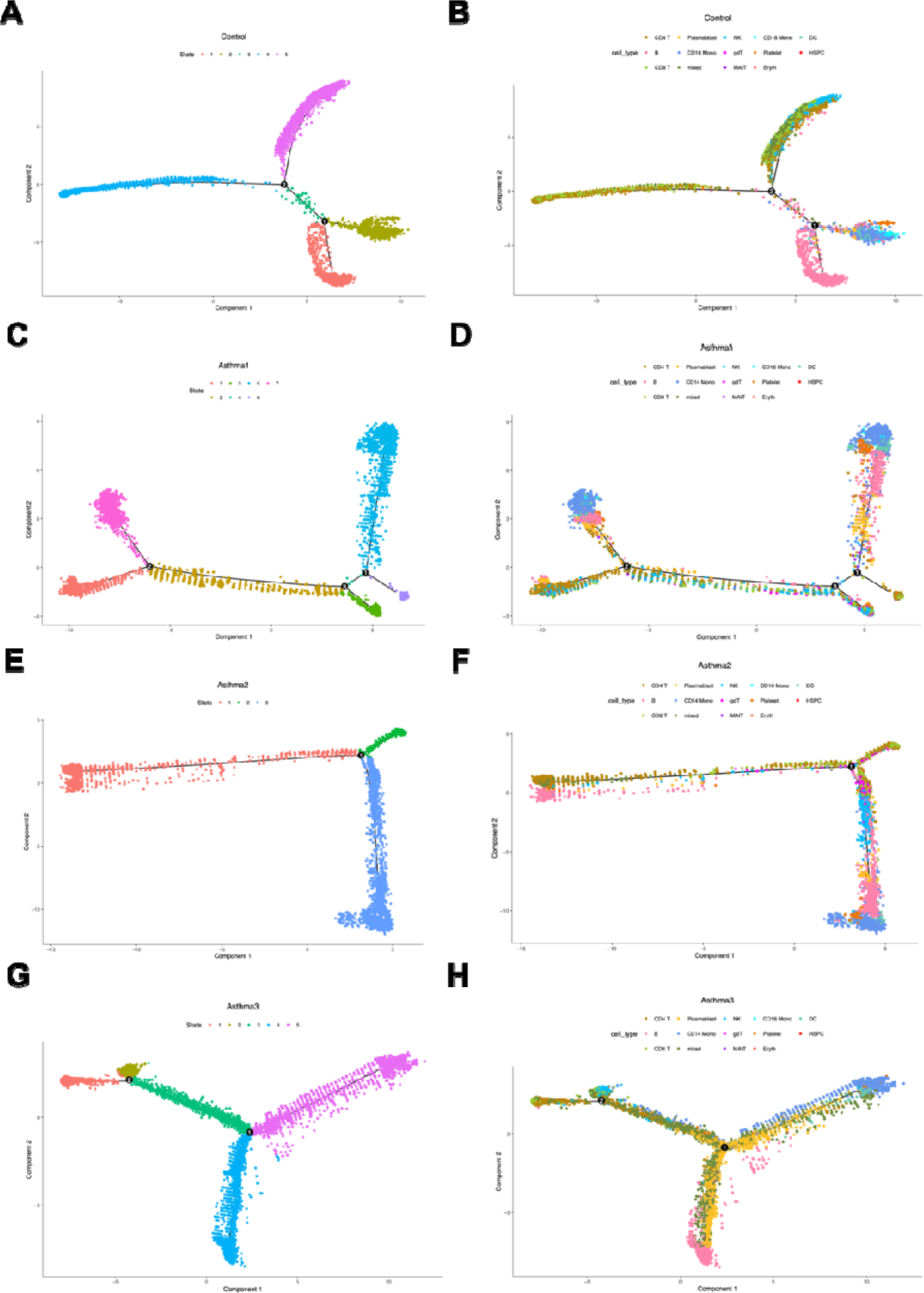
Pseudo-time analysis of all cells in four sample groups. A. The differentiation trajectory of all cells in healthy controls by state. B. The differentiation trajectory of all cells in healthy controls by cell types. C. The differentiation trajectory of all cells in asthma patient 1 (Ast1) by state. D. The differentiation trajectory of all cells in asthma patient 1 (Ast1) by cell types. E. The differentiation trajectory of all cells in asthma patient 2 (Ast2) by state. F. The differentiation trajectory of all cells in asthma patient 2 (Ast2) by cell types. G. The differentiation trajectory of all cells in asthma patient 3 (Ast3) by state. H. The differentiation trajectory of all cells in asthma patient 3 (Ast3) by cell types.

Based on the major cell types and numbers, state 1 in healthy controls can be classified as B lineage, state 2 can be classified as monocyte lineage, state 3 is a mixed lineage (B and T), state 4 can be classified as T lineage, and state 5 can be classified as T-NK lineage (Figure 3b and Supplement Figure 2a). Myeloid lineage (monocytes) and two lymphoid lineages (B cells and T cells) are clearly differentiated in the PBMCs of healthy controls. However, the clear cell development patterns disappear in 3 asthma patients. Each patient exhibits unique cell developmental patterns different from healthy controls. A1 has 7 states and 4 of them are T lineages according to the major cell numbers (state 1, 2, 3, and 6, Figure 5d and Supplement Figure 2b). A2 has only 3 states and a part of B cells are mixed with T cells (state 1, Figure 5f and Supplement Figure 2c). A3 has 5 states, but B cells are mixed with T cells in state 3 (state 1, Figure 5h and Supplement Figure 2d). Although different childhood asthma patient has a different cell developmental state, abnormal lymphoid lineage development is observed in all 3 patients (multiple T lineage states or no clear B lineage state). This result is consistent with the expression feature analyses showing that adaptive immunity is repressed in asthma patients. T cells and B cells are a major constitution of adaptive immune system and their poor development naturally leads to insufficient the adaptive immunity.

### Pseudo-time expression dynamic analyses of proto-oncogenes and inflammatory response related genes

The abnormal cell developmental trajectories in asthma patients should be accompanied by abnormal gene expression patterns. JUN gene is a transcription factor regulated by various extracellular stimuli including peptide growth factors, pro-inflammatory cytokines, and even UV irradiation^12^. SPI1 gene is also a transcription factor regulating gene expression during myeloid and B-lymphoid cell development^13^. Both of them are proto-oncogenes essential for cellular differentiation^14, 15^. We further include six inflammatory response related genes in pseudo-time expression dynamic analyses. Their express dynamics were plotted along cell developmental trajectories in both health control and three asthma patients.

In health controls, we observed the onset expression of JUN in the early pseudo-time state and a peak expression of six inflammatory response related genes in the early-middle pseudo-time state. SPI1 is expressed in the late pseudo-time state (Figure 6a). In A1 and A2, the expression dynamics of JUN, SPI1, and six inflammatory response related genes are almost the same (Figure 6b and 6c). JUN’s onset expression is repressed while SPI1 exhibits an onset expression in the early pseudo-time state. S100A8, S100A9, and S100A12 show an early onset expression rather than a peak expression in the early-middle pseudo-time state. IL7R, IL32, and CCL5 start to express in the early-middle pseudo-time state as health control, but they show a persistent expression level instead of an obvious expression peak. Notably, IL7R, IL32, and CCL5 all participate in the biological process of negative regulation of the T cell apoptotic process (GO:0070233). It implies that the T cells have prolonged cell survival in A1 and A2. A3 shows a different expression dynamic pattern from health control, A1, and A2 (Figure 6d). In A3, JUN is also repressed while the expression patterns of IL7R, IL32, and CCL5 are more similar to health control. S100A8, S100A9, S100A12, and SPI1 show a persistent expression level without decline. Since SPI1 activates gene expression during myeloid and B-lymphoid cell development and S100 protein family plays a role in cell growth and differentiation, their persistent expression pattern indicates an abnormal lymphoid cell development fate in A3.

**Figure 6.**
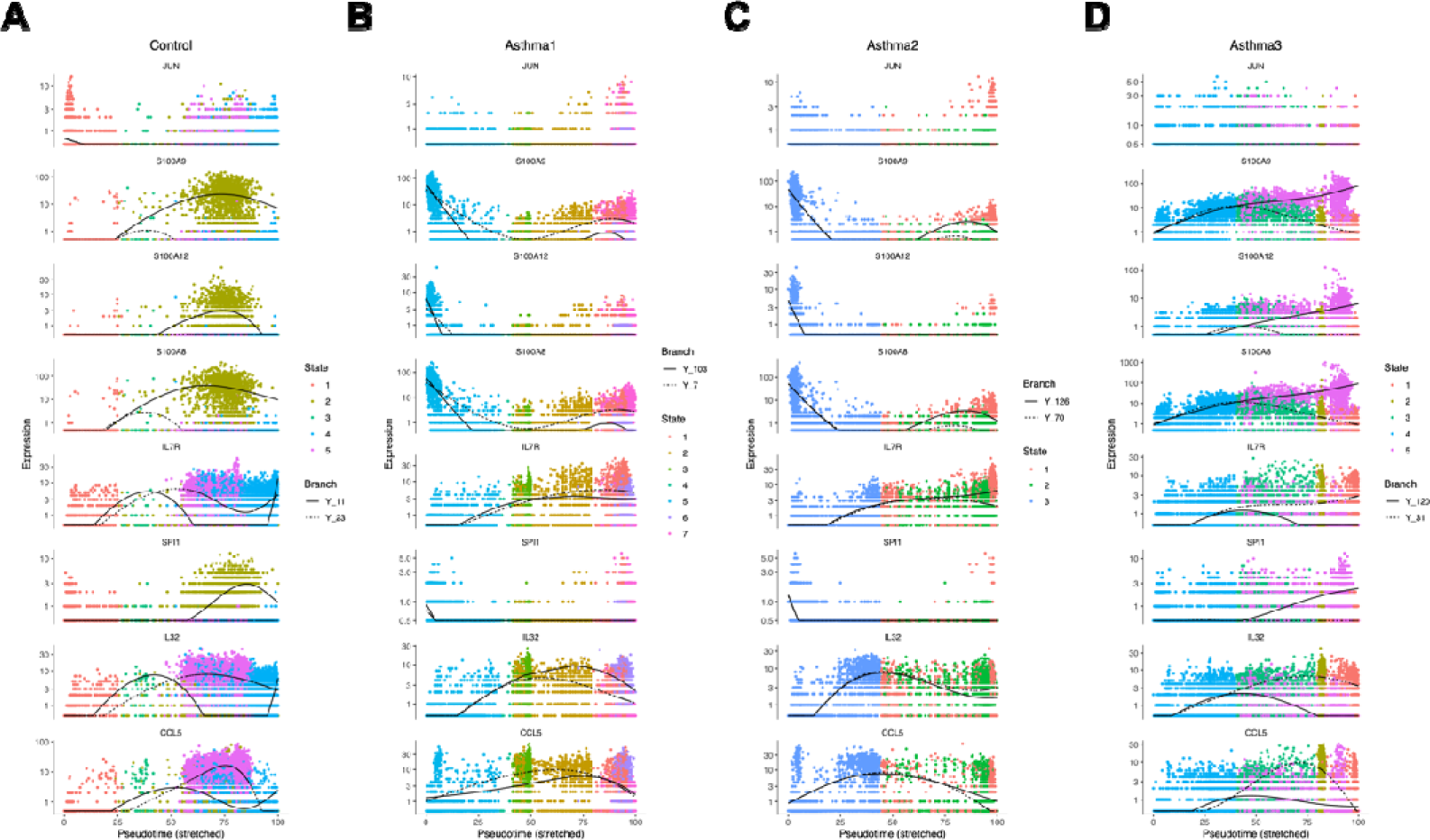
Pseudo-time analyses of expression dynamics of two cell-fate and six inflammation related genes in four sample groups. A. Expression dynamics two cell-fate and six inflammation related genes in health control. B. Expression dynamics two cell-fate and six inflammation related genes in health control in asthma patient 1 (Ast1). C. Expression dynamics two cell-fate and six inflammation related genes in health control in asthma patient 2 (Ast2). D. Expression dynamics two cell-fate and six inflammation related genes in health control in asthma patient 3 (Ast3).

### DCs function as a signaling hub in cell communication and play a role in asthma heterogeneity

We performed cell communication analyses for both health control and asthma patients to investigate their cell-cell interactions. The number of interactions and interaction strength for health control and three asthma patients are shown in Figure 7. Generally, asthma patients have a weaker interaction strength than health control and A3 has the smallest number of interactions and the weakest interaction strength among four samples. Among the major immune cell types, we find that dendritic cells have the largest number of both incoming and outgoing interaction numbers(Supplemental Figure 3a, 3b, 3c, and 3d). DCs are known to be the messenger cells communicating between innate and adaptive immune systems^16^. They are also known as the antigen presenting cells which interact with T cells and B cells to activate and regulate the adaptive immune response^17^. Although DCs don’t have the strongest interaction strength among the major immune cell types (Supplemental Figure 4a, 4b, 4c, and 4d), they surely function as a signaling hub in cell communication based on cell communication analyses.

**Figure 7.**
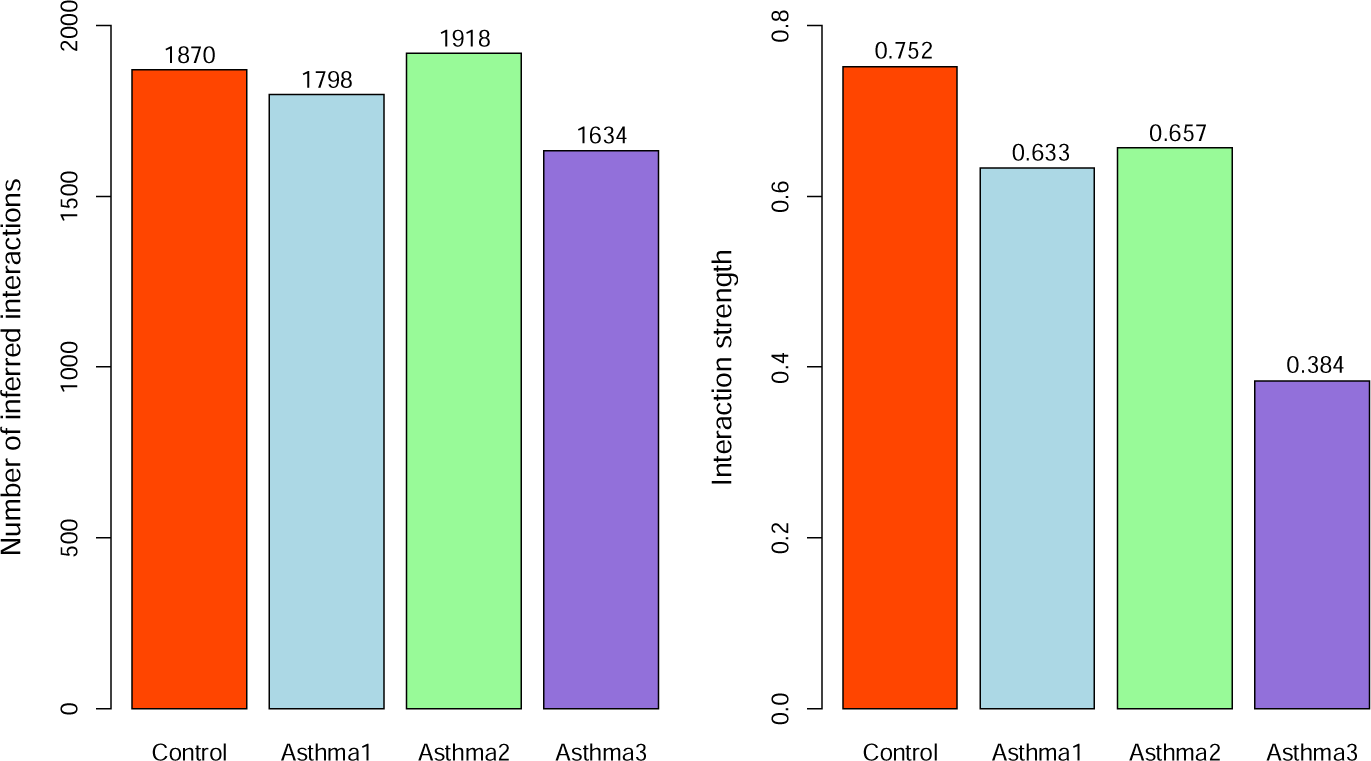
The number of cell-cell interactions and interaction strength for four sample groups.

We further examined the DCs’ signaling differences between health control and asthma patients. Using health control as a signaling background, the signaling changes were detected in three asthma patients. The DCs of A1 has a specific incoming signal of annexin (Supplemental Figure 5a). The DCs of A2 has a specific outgoing signal of resistin (Supplemental Figure 5b). The DCs of A3 has a specific incoming signal of annexin and a specific outgoing signal of resistin (Supplemental Figure 5c). Annexin A1, A2 and A5 have a role in the regulation of inflammation activation and may serve as the potential biomarkers for asthma^18–20^. They all show an elevated expression tendency in asthma cases (Supplemental Figure 6). Resistin also serves a potential biomarker for asthma and is related to inflammation as well^21, 22^. Our results show that annexin and resistin are actually two opposite directional signals for the DCs of asthma patients. One of them is a sufficient predictor of asthma and both of them indicate a severe symptom of asthma, since A3 is an acute severe asthma patient. Both annexin and resistin show a self-reinforce trend in the CD14 monocytes of asthma patients (Figure 8a and 8b). The upregulated signaling interactions in asthma are mainly centered on the molecular component of external side of plasma membrane and cell adhesion molecules (GO:0009897 and hsa04514, Supplemental Figure 7a, 7b, and 7c). Furthermore, we found that CLEC2B-KLRB1 and LGALS9-CD44 signaling interactions are downregulated in all three asthma patients (Supplemental Figure 8a, 8b, and 8c). CLEC2B-KLRB1 interaction mediates the activation of NK cells and monocytes^23^. LGALS9-CD44 interaction enhances the stability and function of adaptive regulatory T cells^24^. The down-regulated CLEC2B-KLRB1 and LGALS9-CD44 interactions proposes a weakened functions of NK cells, monocytes and T cells in asthma patients.

**Figure 8.**
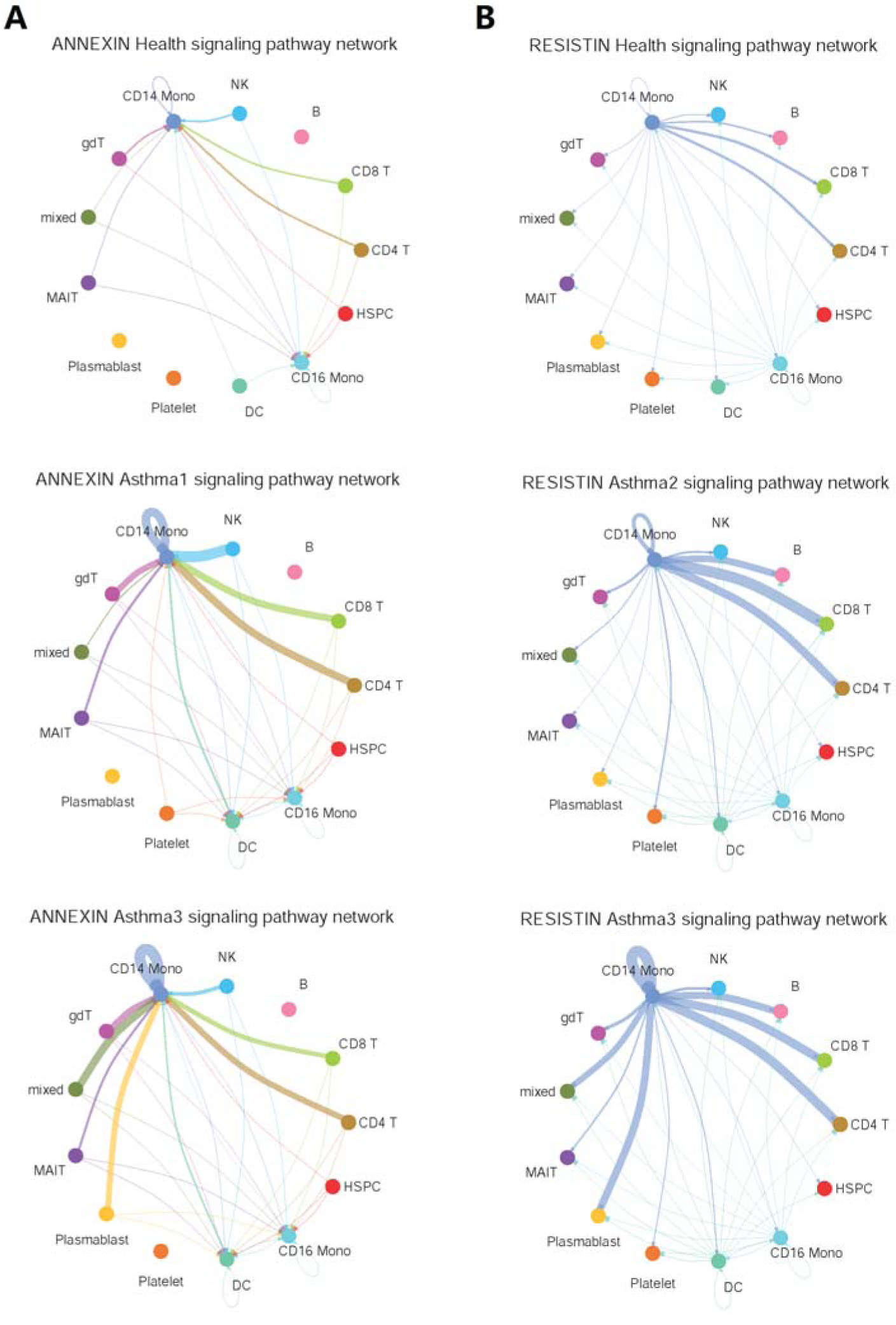
The annexin and resistin signaling pathway network of four sample groups. The annexin signaling pathway network is shown for health control, asthma patient 1 (Ast1) and asthma patient 3 (Ast3). The resistin signaling pathway network is shown for health control, asthma patient 1 (Ast1) and asthma patient 2 (Ast2).

### Plasma blast cells in A3 express T cell receptor genes

Patient A3, a severe asthma sufferer who has been admitted to the pediatric intensive care unit, exhibits a unique group of cells not present in healthy controls A1 and A2 (Figure 1a). This group predominantly consists of plasma blast cells along with a mixed cluster comprising plasma blast cells (35.2%), erythrocytes (26.5%), CD14 monocytes (14.4%), CD4 T cells (11.2%), CD8 T cells (7.7%), and other cells (5%).

Upon comparing the gene expression profile of A3’s plasma blast cells with those of the healthy controls, A1 and A2, we discovered an unexpected finding: the plasma blast cells in A3 express a range of T cell receptor genes, including TRBV20-1, in addition to immunoglobulins (Figure 9a). Further investigation into the expression levels of classical markers for plasma blast cells (CD38, CD19, CD27), CD4 T cells (CD8A, CD27), CD8 T cells (CD40LG, CD27), and NK cells (GNLY, NKG7) revealed through a violin plot that all these markers are indeed expressed in A3’s plasma blast cells (Figure 9b). This suggests that the plasma blast cells in A3 likely possess dual cellular identities, exhibiting characteristics of both plasma blast cells and T cells, which accounts for the expression of T cell receptor genes.

**Figure 9.**
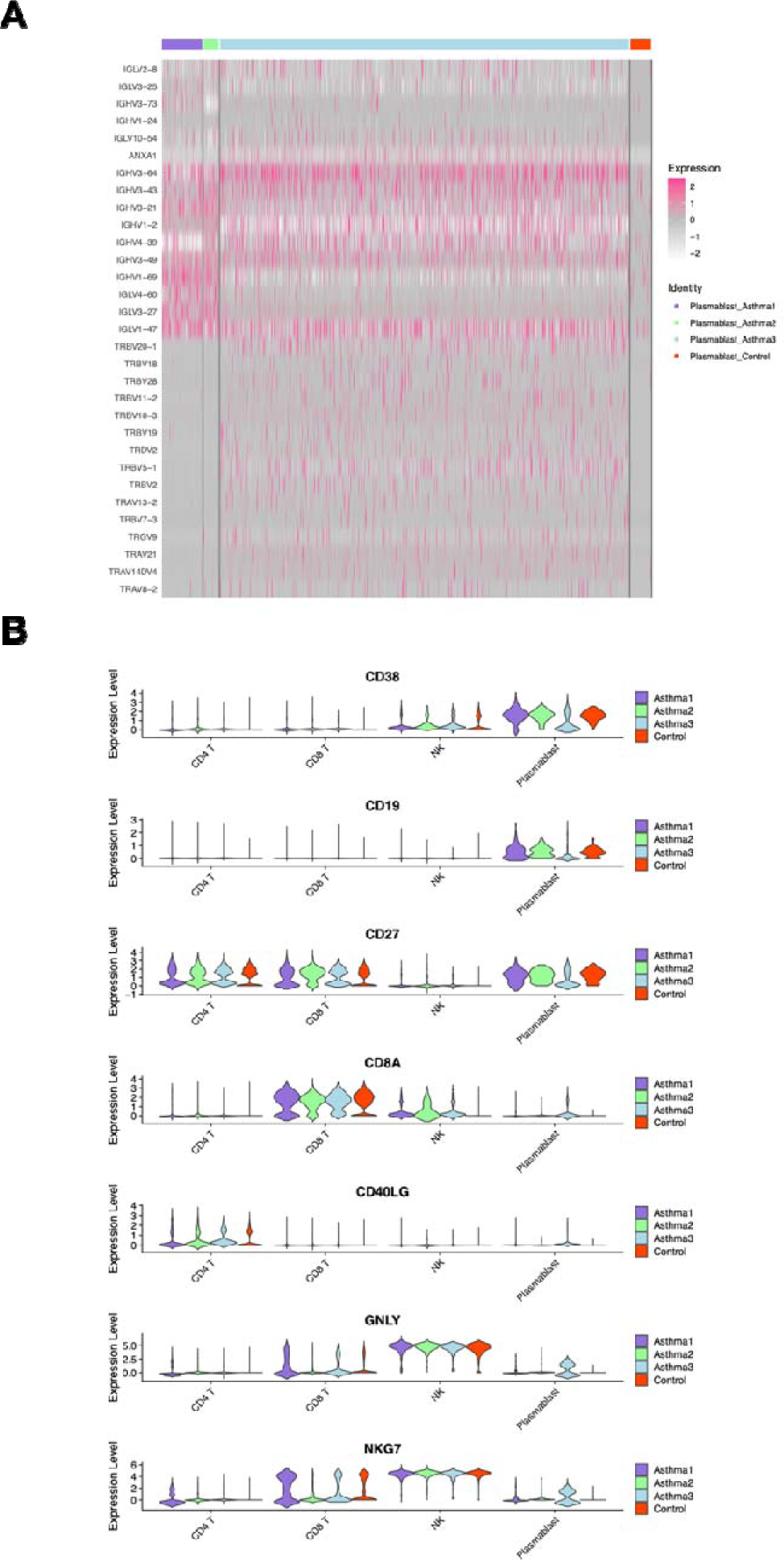
The dual cell identity of the plasma blast cells in asthma patient 3. A. The expression heatmap of immunoglobulin (IGLV or IGHV) genes and T cell receptor (TRBV or TRAV) genes in the plasma blast cells of four sample groups. B. The violin plot of CD38, CD19, CD27, CD8A, CD40LG, GNLY and NKG7 gene expression levels across CD4+ T cell, CD8+ T cell, NK cell and plasma blast cells among four sample groups. CD38 and CD19 for the markers for plasma blast cells. CD27 is the marker for both plasma blast cells and T cells. CD8A, CD40LG, GNLY and NKG7 are the markers for T cells.

### The expression level of RETN in lymphoid linage cells

The feature plot shows that the RETN expression is not evenly distributed among different cell clusters. CD14 mono, CD16 mono, and dendritic cells have an especially high expression level of RETN. The violin plot further shows that RETN expression level in these four clusters is always higher in 3 asthma cases than in healthy controls. RETN encodes resistin, then we tested the level of resistin in plasma between children with asthma and healthy controls, we found that resistin in the serum increased significantly in the asthma group than in controls (Figure 10).

**Figure 10.**
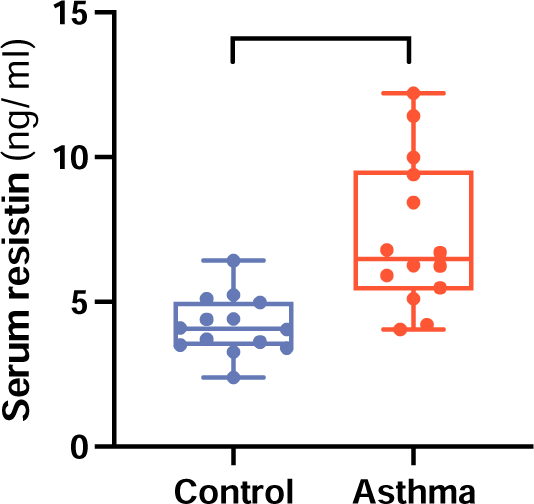
The expression of RETN gene in four sample groups. The level of serum resistin between asthma patients and health controls(\*\*\**p* < 0.001).

## Discussions

In individuals with allergic airway disease, such as allergic asthma, the airways are already sensitized to specific allergens. This means that the immune system has developed an exaggerated response to these allergens, leading to chronic inflammation and airway hyperresponsiveness. During an asthma exacerbation, this preexisting allergic airway disease is further aggravated by additional inflammatory stimuli or triggers. Nevertheless, the etiology of childhood asthma is so far poorly understood^25^.

The analyses of PBMCs from three childhood asthma patients show that childhood asthma is a complex disease with distinct individualities at the cellular level. The overall expression features among three asthma patients’ PBMCs demonstrate that innate immunity is commonly upregulated while adaptive immunity is commonly downregulated in childhood asthma, but each patient has different molecular phenotypes. The common upregulated genes in the HSPCs of asthma patient are related to the cellular components of extracellular exosome and ER to Golgi transport vesicle membrane and the common downregulated genes in the HSPCs of asthma patients are related to S100 protein binding. The specific downregulated genes in the HSPCs of each asthma patients converge on immunity and T cell receptor related biological entities. Because HSPCs give the rise to the other cells, they are a cellular source of immunity. The expression abnormality in HSPCs would surely influence their descendant cells. The downregulation of S100 protein binding in the HSPCs indicates that S100 proteins are in an active form in the HSPCs of asthma patients. The downregulation of immunity related genes in HSPCs is a sign of hypo-immune tendency started from the cellular source in asthma cases. The active S100 protein and hyo-immune tendency in the HSPCs of asthma patients seem to lead to over innate immunity and low adaptive immunity for all cells of asthma patients. Although childhood asthma has common pathophysiological features at the cellular level, there are more specific differentially expressed genes (DEGs) in each patient than common DEGs, which proposes that the genetic causes of asthma vary across different children or different allergens can trigger different immune genes in a pediatric patient’s immune system. These two explanations are mutually inclusive. Future studies includes bone marrow samples might give us much clearer views for this question, since HSPCs are generated from bone marrow.

The single-cell trajectory analyses further demonstrate the cell developmental heterogeneities among asthma patients. Different asthma patient has different developmental states. The abnormal lymphoid lineage development is observed in all 3 patients. A1 has multiple T cell states while A2 and A3 have a poorly developed lymphoid state of mixed B and T cells. The expression of T cell receptor genes is repressed in the HSPCs of all three asthma patients, although this repression is not statically significant in A2 and A3. It could partly explain the abnormal cell developmental states in asthma patients and the developmental heterogeneities among them. The delayed expression of T cell receptor genes could lead to the HSPC’s descendant cells having difficulty receiving the lymphoid development signals. In A1, such deficiency is responded to the development of multiple T cell states by A1’s immune system, which seems to compensate for the lack of a proper T cell developmental state. In A2 and A3, the delayed expression of T cell receptor genes is less severe than that in A1, which leads to the underdevelopment of lymphoid lineage. The abnormal single-cell developmental trajectories in asthma patients are also accompanied by the key genes’ aberrant expression dynamics. JUN and SPI1 are two proto-oncogenes, both of which have multiple functions in cell fate decisions^15, 26^. The expression of JUN is repressed in the early stage of cell trajectories for all three asthma patients. It indicates that the cell developmental fate of asthma patients has trouble at the very beginning, which is consistent with the expression abnormality in the HSPCs of asthma patients. In A1 and A2, the repression of JUN seems to be replaced by the expression of SPI1. In A3, the beginning of the expression of SPI1 is consistent with that in health control, but the expression of SPI1 shows no decline like in health control. It implies that the developmental fates of distal cells in A3 could be influenced by SPI1.

The cell-cell communication analyses further revealed the possible mechanisms underlying the etiology of asthma. Although the number of cell-cell interactions is similar in health control and asthma cases, the strength of cell-cell interactions is much weaker in asthma patients, especially in A3. The overall weak signaling strength is a clear sign of cell-cell communication disorder in asthma. It is in accordance with the abnormal cell developmental trajectories of asthma. DCs induce primary immune responses by processing and presenting antigen material on the cell surface of T cells^27^. They function as the signaling hub communicating between innate and adaptive immune systems. Two aberrant signals, annexin, and resistin, are found for the DCs of asthma, both of which play a part in inflammatory response. Inflammation is a generic immune response and is considered as a mechanism of innate immunity. It explains why innate immunity is hyperactive in asthma patients. In our cases, each asthma patient has his or her aberrant signaling pattern, annexin for A1, resistin for A2, and both annexin and resistin for A3. It further explains the heterogeneities among asthma patients. Resistin as an outgoing signal from DCs has been reported as a predictor of asthma^21^. Our ELISA results show that its expression can be detected in 12 enlisted childhood asthma patients.

A3 is a special case in our study for the patient with severe symptoms compared to A1 and A2 whose symptoms are mild. Severe asthma means strong inflammation in patient’s airway which further causes breathlessness and continuous cough. The profiling of A3’s PBMCs first presents the patient’s severity in our study. A3 has two extra cell clusters compared with health control, A1 and A2. These two extra cell clusters are annotated as mixed cells and plasma blast cells. The UMAP plot shows that the mixed cells and plasma blast cells in A3 are stemmed from CD4 T cells. The expression features of A3’s plasma blast cells demonstrate that they express T cell receptor genes as well. It is noticed that SPI1 has a persistent expression dynamic in A3. That A3’s plasma blast cells show a duality of T cell identity could be linked to the SPI1’s aberrant expression pattern. SPI1 might influence the developmental fate of plasma blast cells in A3 and further exacerbate the patient’s symptoms.

## Conclusions

Our research substantiates that childhood asthma is a multifaceted disease characterized by an upregulation of the innate immune system and a downregulation of the adaptive immune response. Single-cell RNA profiling of PBMCs from asthma patients has uncovered a variety of expression patterns among HSPCs, along with aberrant cell developmental trajectories and diminished cell-cell interaction signaling. While significant heterogeneity exists among different cases of asthma, the pathological features converge on certain commonalities, particularly the abnormal development of the lymphoid lineage and an overabundance of expression in genes related to S100 protein binding and immune response. Notably, S100A8, S100A9, S100A12, and RETN, along with ANXA1, ANXA2, and ANXA5, have emerged as pivotal molecular markers in asthma, directly implicated in the inflammatory response.

## Data Availability

All data produced in the present study are available upon reasonable request to the authors

## Contributions

Xiaoyan Dong and Guohui Ding designed the study. Danying Zhu wrote the manuscript. Libing Shen analysis the data. Zeyu Zeng did literature search. Lang Yuan and Na Dong contributed clinical samples. Chao Wang and Ming Chen collected the clincal data. Lijian Xie and Xiaoyan Dong funded the single cell sequencing.

## Acknowledgements

This study was funded by Jinshan Distinct key medical specialty project (JSZK2023A04) and Institute of Pediatric Infection, Immunity, and Critical Care Medicine Cultivate Research Project.

## Data Sharing Statement

All data generated or analyzed during this study are included in this article. Further enquiries can be directed to the corresponding author.

## Supplemental Figures

**Supplemental Figure 1.**
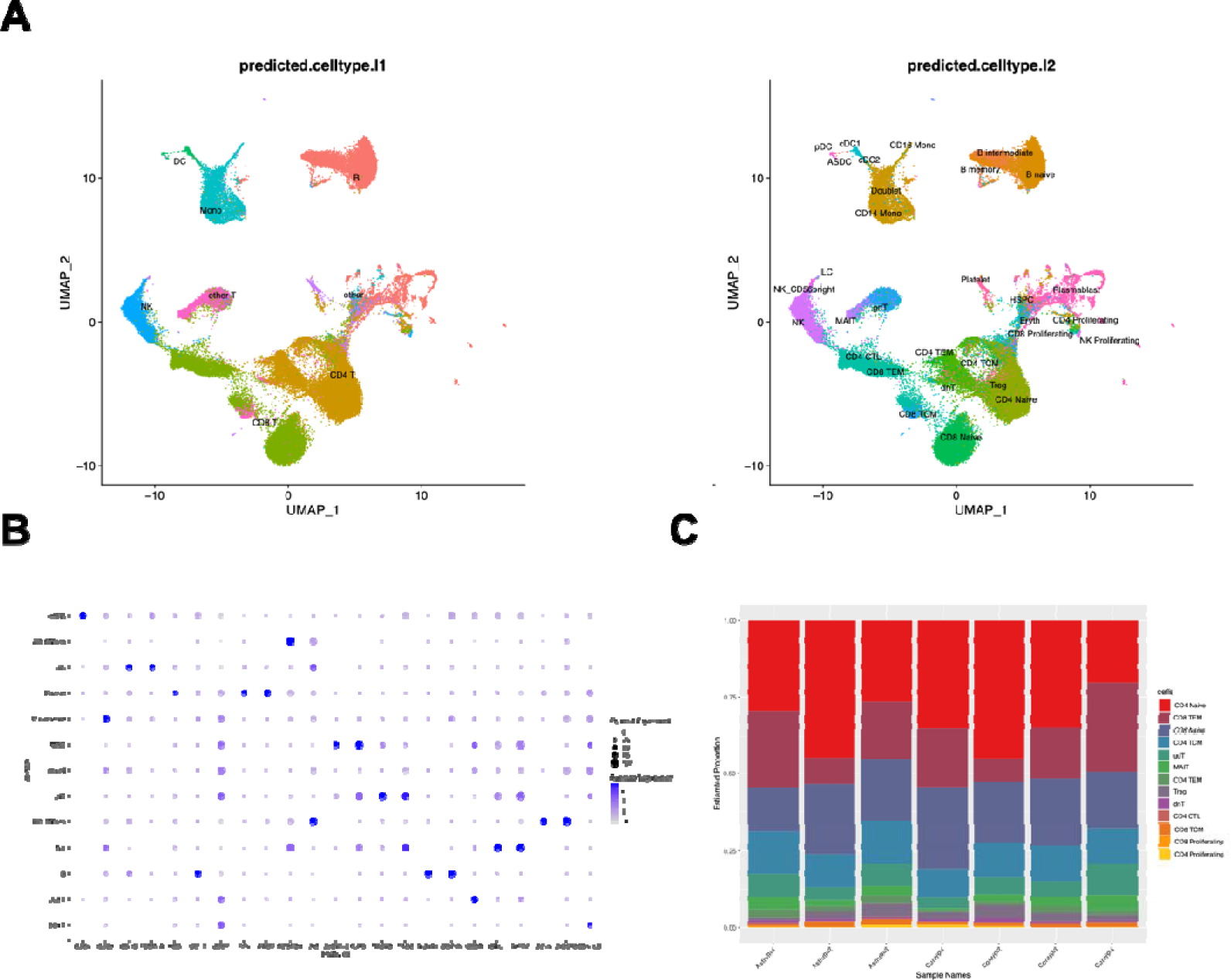
Integrated single-cell profiling of PBMCs for four sample groups. A. The predicted cell types for the integrated single-cell profiling of PBMCs for four sample groups B. Expression of canonical gene markers for each cell type in four sample groups based on integration analysis. C. The detailed classification and proportion of T cells for all seven samples in our study.

**Supplemental Figure 2.**
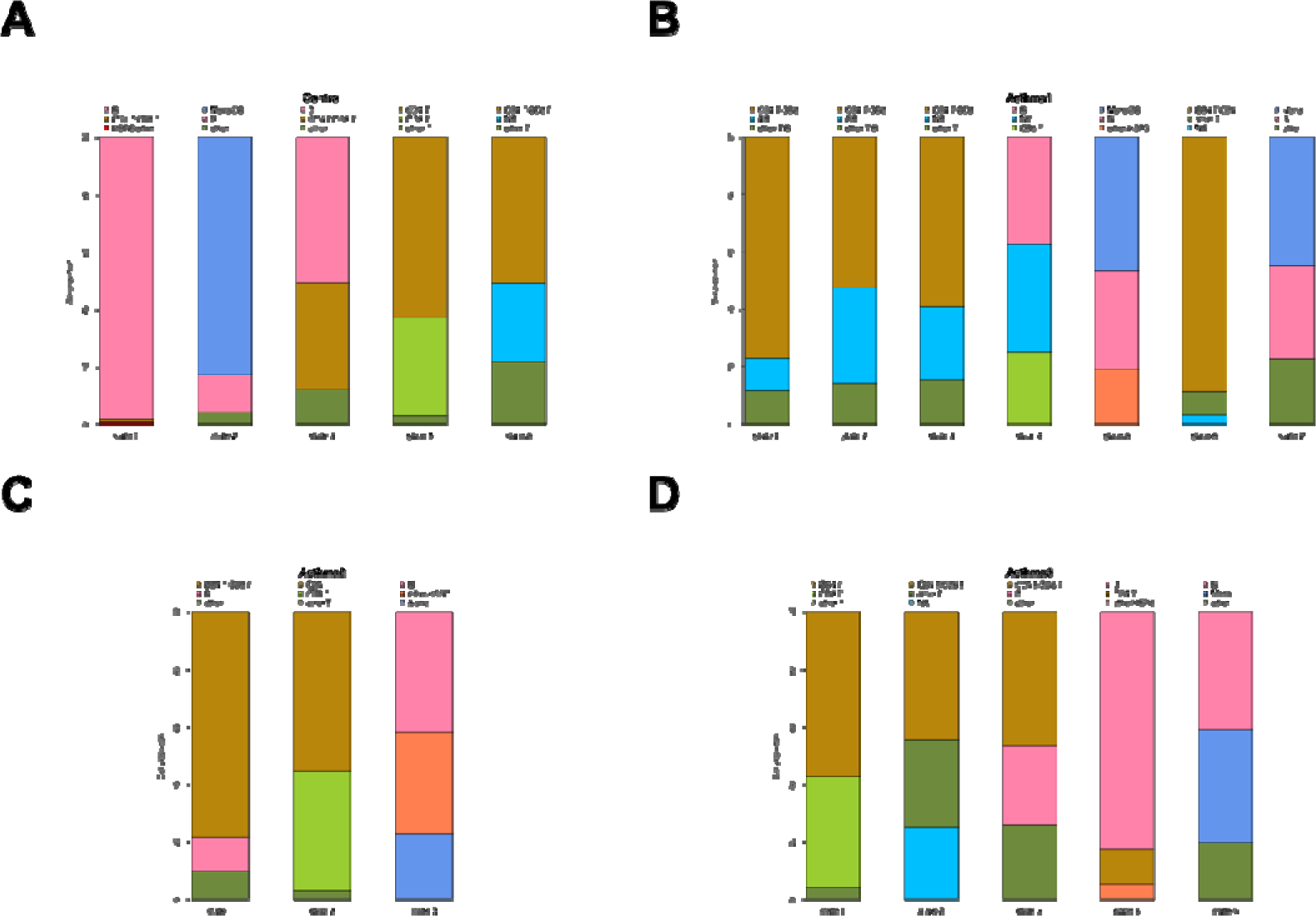
Pseudo-time analysis of all cells in four sample groups. A. The proportion of major cell types in each state for healthy controls. B. The proportion of major cell type in each state for asthma patient 1 (Ast1). C. The proportion of major cell type in each state for asthma patient 2 (Ast2). D. The proportion of major cell type in each state for asthma patient 3 (Ast3).

**Supplemental Figure 3.**
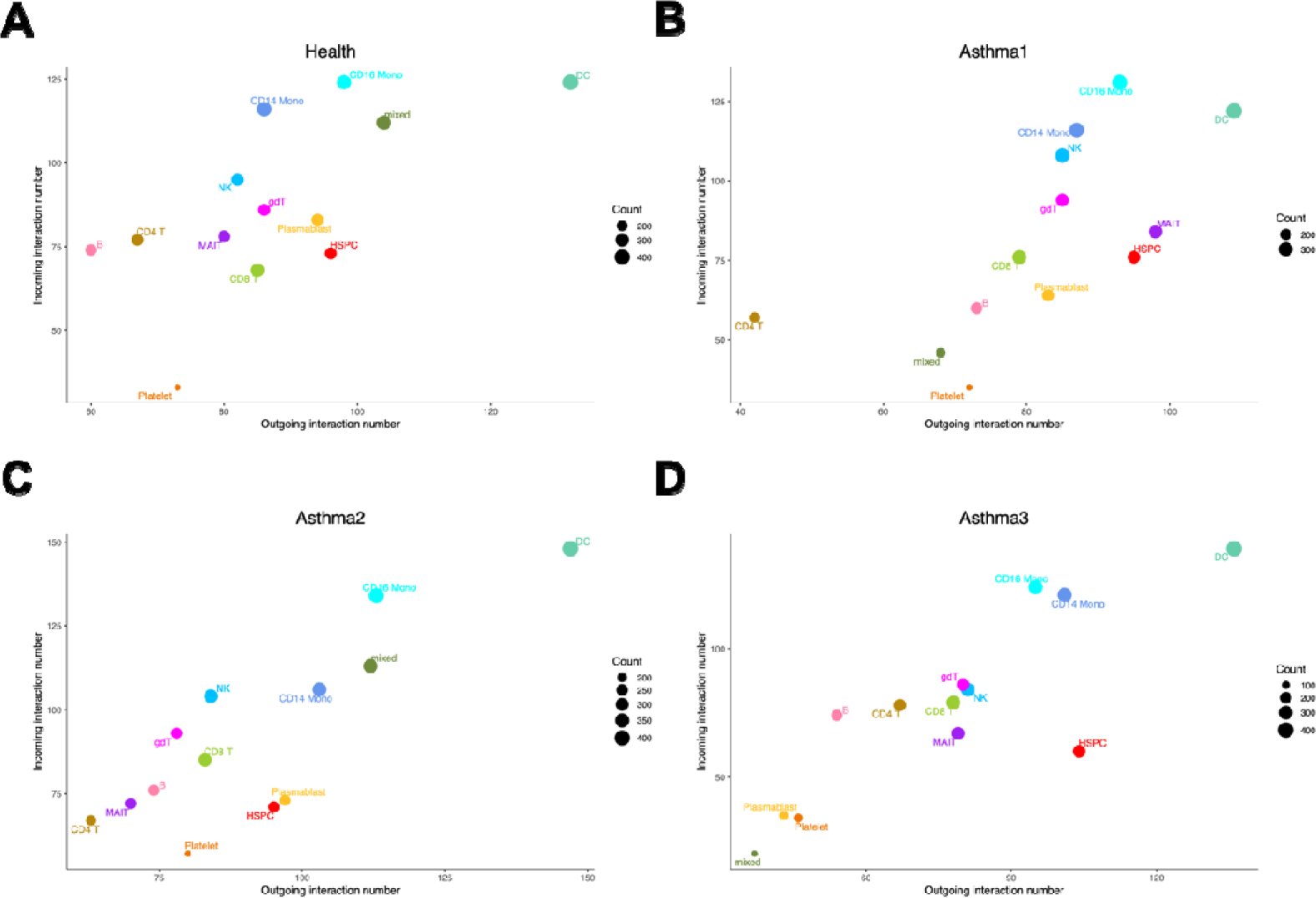
The number of incoming and outgoing interactions for each cell type in four sample groups. A. The number of incoming and outgoing interactions in health control. B. The number of incoming and outgoing interactions in asthma patient 1 (Ast1). C. The number of incoming and outgoing interactions in asthma patient 2 (Ast2). D. The number of incoming and outgoing interactions in asthma patient 3 (Ast3).

**Supplemental Figure 4.**
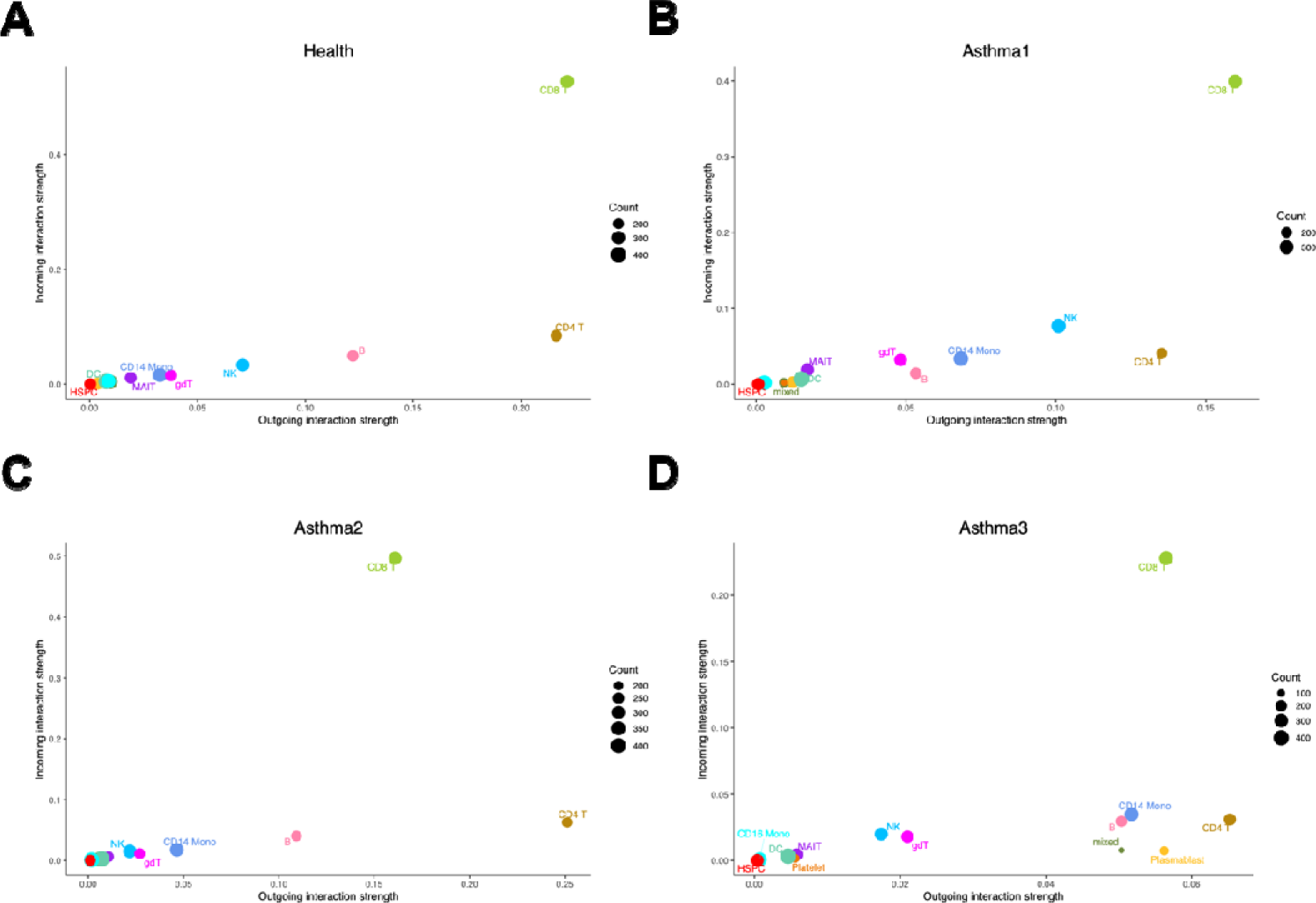
The number of incoming and outgoing interaction strength for each cell type in four sample groups. A. The number of incoming and outgoing interaction strength in health control. B. The number of incoming and outgoing interaction strength in asthma patient 1 (Ast1). C. The number of incoming and outgoing interaction strength in asthma patient 2 (Ast2). D. The number of incoming and outgoing interaction strength in asthma patient 3 (Ast3).

**Supplemental Figure 5.**
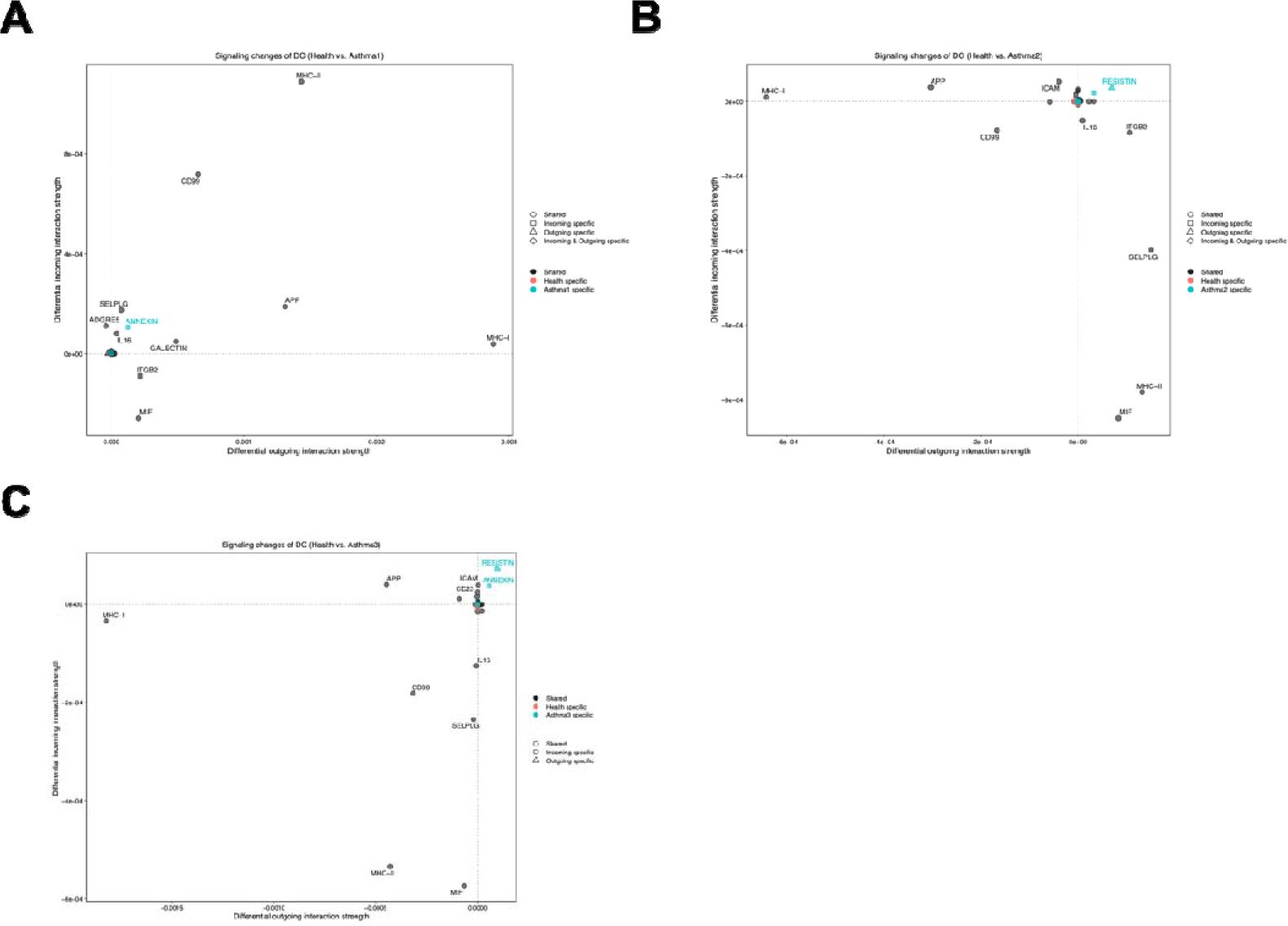
The signaling changes of DCs in three asthma patients (health control as the backgroup). A. The signaling changes of DCs in asthma patient 1 (Ast1). B. The signaling changes of DCs in asthma patient 2 (Ast2). C. The signaling changes of DCs in asthma patient 3 (Ast3).

**Supplemental Figure 6.**
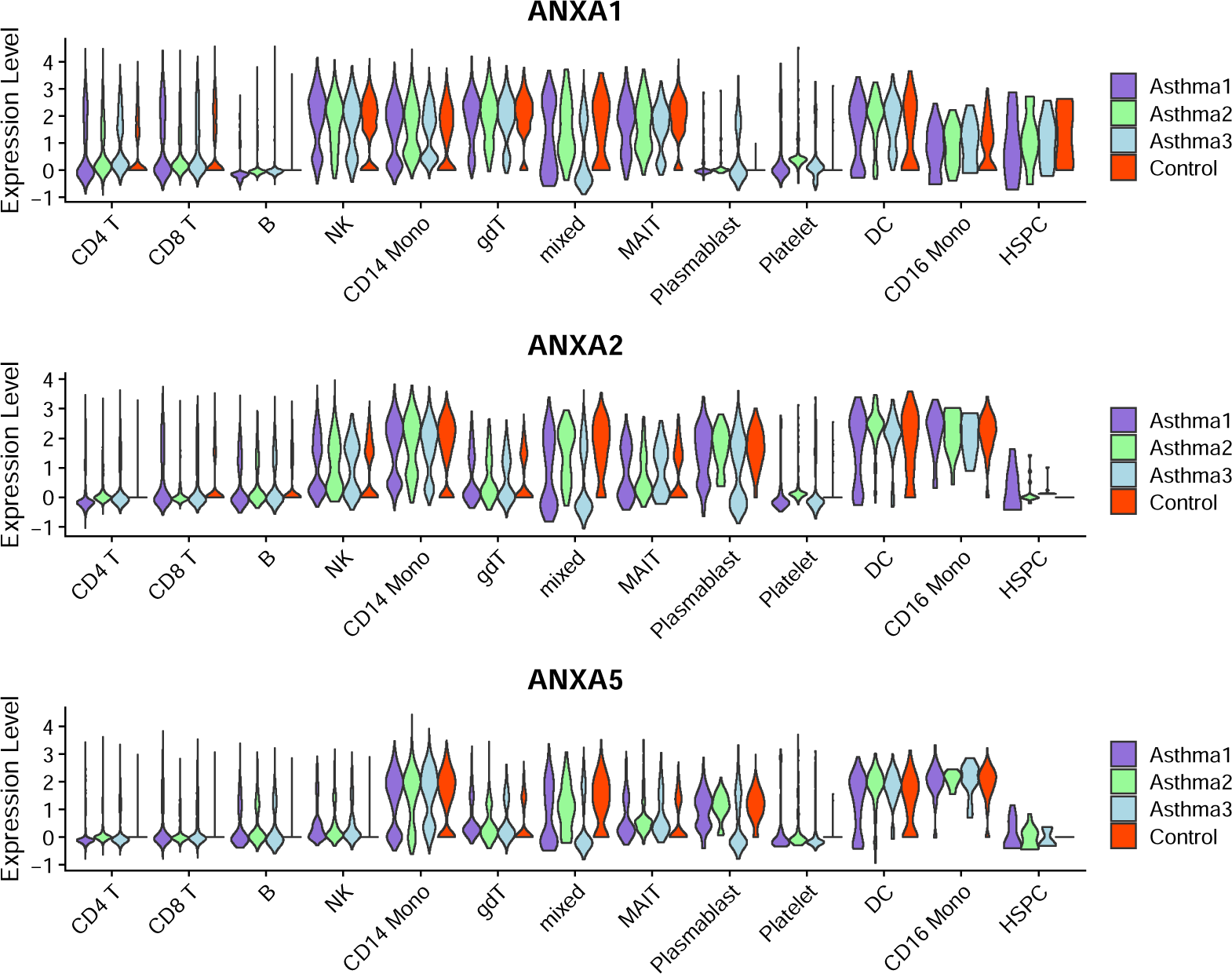
The violin plot of ANXA1, ANXA2 and ANXA2 gene expression levels across different cell types for four sample groups.

**Supplemental Figure 7.**
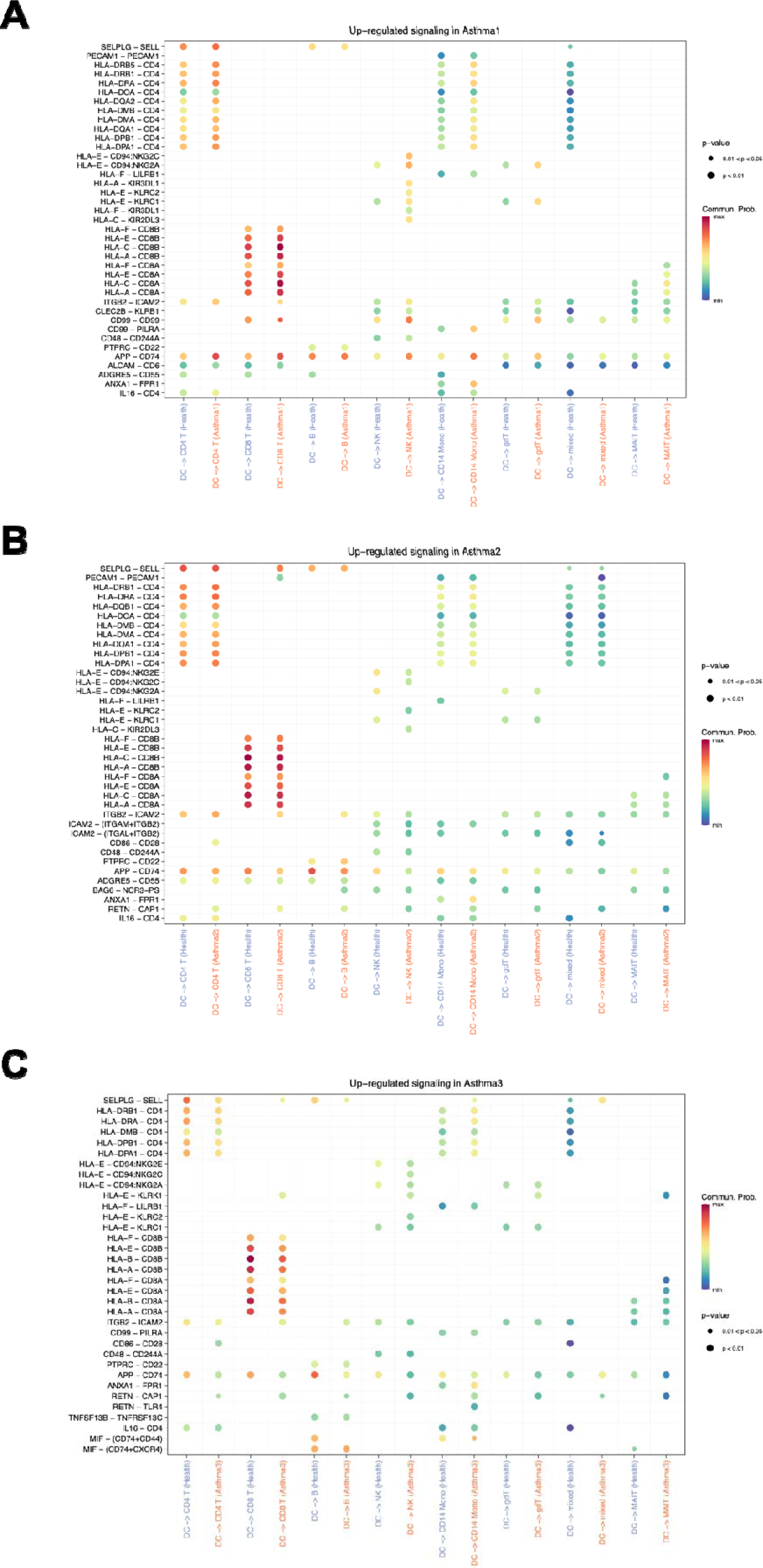
The upregulated signaling from DCs to the other cell types in three asthma patients. A. The upregulated signaling from DCs to the other cell types in asthma patient 1 (Ast1). B. The upregulated signaling from DCs to the other cell types in asthma patient 2 (Ast2). C. The upregulated signaling from DCs to the other cell types in asthma patient 3 (Ast3).

**Supplemental Figure 8.**
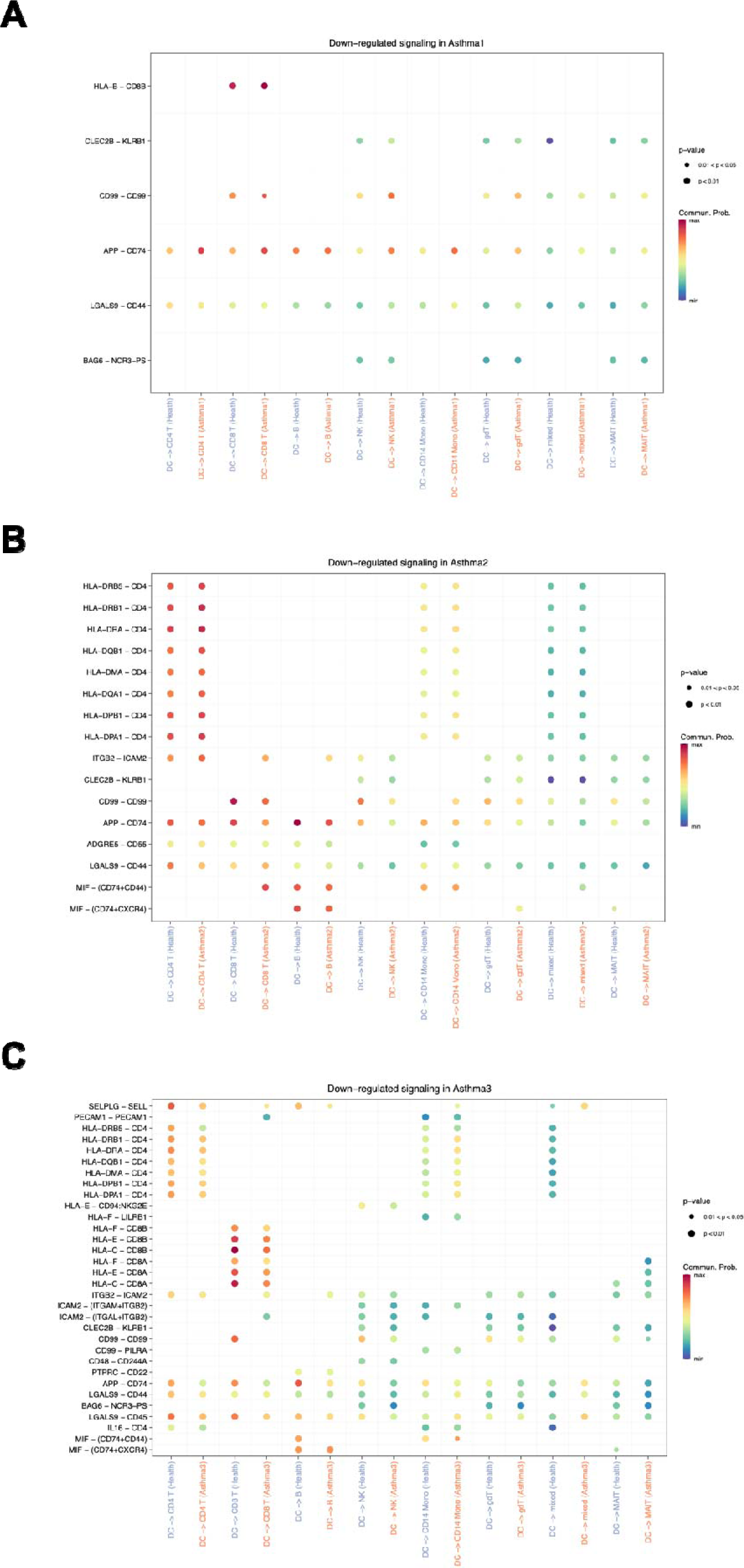
The downregulated signaling from DCs to the other cell types in three asthma patients. A. The downregulated signaling from DCs to the other cell types in asthma patient 1 (Ast1). B. The downregulated signaling from DCs to the other cell types in asthma patient 2 (Ast2). C. The downregulated signaling from DCs to the other cell types in asthma patient 3 (Ast3).

